# Integrated analyses of longitudinal trends of antibiotic-resistant bacteria in wastewater, clinical resistance data, and antibiotic consumption in Switzerland

**DOI:** 10.1101/2025.11.22.25340789

**Authors:** Sheena Conforti, Melissa Pitton, Patrick Schmidhalter, Anna Wettlauffer, Catherine Plüss-Suard, Andreas Kronenberg, Timothy R. Julian

## Abstract

Antimicrobial resistance (AMR) surveillance requires approaches that monitor both clinical and community-level dynamics. We monitored antibiotic-resistant bacteria in Swiss wastewater and compared these results with human resistance and antibiotic consumption data from the national surveillance network ANRESIS. Between 2021 and 2024, 772 samples from six wastewater treatment plants were analyzed for *Escherichia coli*, extended-spectrum β-lactamase-producing (ESBL)-*E.coli*, carbapenem-resistant *E. coli* (CR-*E.coli*), *Enterococcus faecium/faecalis*, and vancomycin-resistant enterococci (VRE). The rank order of the proportion of resistance was conserved between wastewater and clinical data. Mean (±standard deviation) wastewater resistance percentages were 2.2±0.8 for ESBL-*E.coli*, 0.4±0.6 for VRE, and 0.1±0.1 for CR-*E.coli*. Clinical resistance percentages were 9.8±0.8 for ESBL-*E.coli*, 2.9±1.4 for VRE, and 0.3±0.1 for CR-*E.coli*. Both datasets showed similar rising trends for ESBL-*E. coli* and VRE, while CR-*E. coli* remained stable in wastewater but increased slightly in clinics. No consistent lead-lag relationships were observed between wastewater resistance, clinical resistance, or antibiotic use, indicating independent short-term dynamics. Resistance percentages in wastewater were not associated with antibiotic use data for either the antibiotics used for treatment or the ones that are selective for the target. These results suggest that wastewater monitoring reflects long-term population-level AMR dynamics, aligning with clinical trends over years but not months.

**Graphical abstract:** 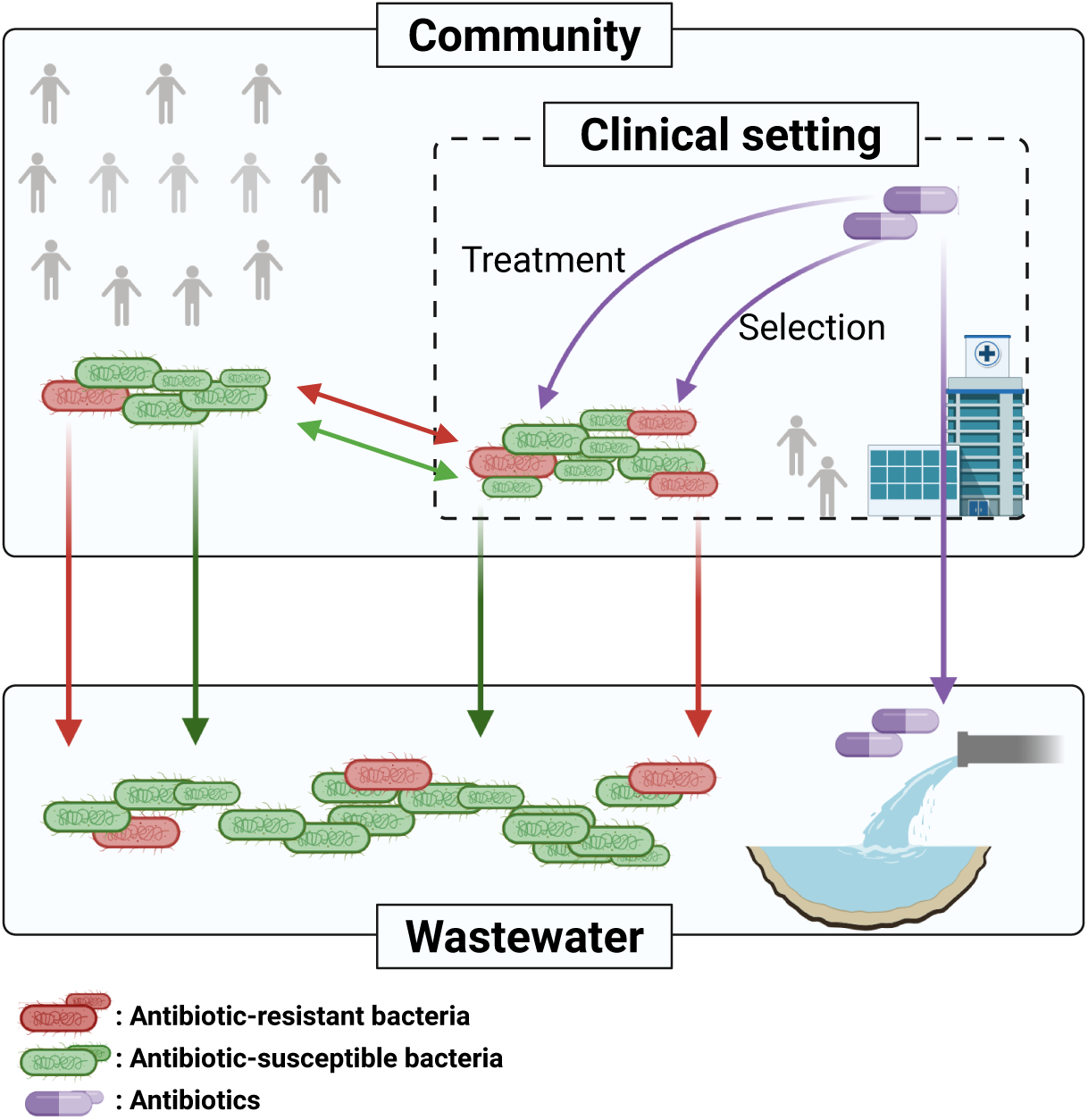

**Synopsis:** Antibiotic-resistant bacteria in Swiss wastewater alongside hospital resistance and antibiotic use data reveal increasing community resistance and show how environmental surveillance can complement clinical surveillance in understanding antibiotic resistance spread.

## Introduction

Antimicrobial resistance (AMR) is a leading global health threat, responsible for an estimated 1.3 million deaths in 2019 (1). In Europe, 25,000 patients die annually from drug-resistant infections (2), and in Switzerland, AMR was associated with 7,160 infections and 276 deaths in 2015 (3). AMR surveillance relies heavily on clinical data, which has multiple limitations. First, clinical surveillance only includes a small fraction of people who visit clinics, does not include information on asymptomatic carriers, and is biased by the availability and prioritization of resources for diagnostics (4,5). Underreporting may be another concern, as not all AMR cases are documented or reported to public health authorities (6). In Switzerland, AMR surveillance is coordinated through the Swiss Centre for Antibiotic Resistance (ANRESIS), which compiles data on resistance and antibiotic consumption from in-and outpatient settings (https://www.anresis.ch, accessed 21 March 2025). ANRESIS is representative for Switzerland, covering 90% of hospital isolates. However, reporting resistance data to ANRESIS is not mandatory, and participation varies across institutions, limiting the completeness of the data.

AMR is not confined to clinical settings, and surveillance focused solely on clinical data provides an incomplete picture, particularly as asymptomatic community carriage rates are high and act as potential sources for clinical AMR infections (7). Measuring AMR in people beyond clinical cases remains difficult, as representative, population-wide screening is expensive and not feasible. As a result, recent research has suggested wastewater may provide a proxy for community-level carriage, using it to estimate AMR trends across populations (8). Previous studies have demonstrated that antibiotic-resistant bacteria (ARB) and antibiotic-resistant genes (ARGs) are excreted in bodily fluids and enter the sewage system, and that these bacteria and genes are detectable in wastewater influent, suggesting wastewater is a reliable proxy for population-level AMR prevalence (9–11). Through cross-sectional analyses, it has been shown that wastewater can elucidate risk factors for AMR, including international travel and social vulnerability (12). Wastewater-based surveillance (WBS) can also provide timely, spatially resolved insights that are independent of healthcare access or clinical reporting behavior. Additionally, WBS is cost-effective (13).

Across Europe, the prevalence of ARB detected in wastewater has shown strong concordance with clinical surveillance data (14). For instance, ARB levels in Nordic wastewater closely mirror regional clinical findings (15), and resistance profiles in Swedish hospital sewage correlate with those from patient urine and blood samples (11). In Switzerland, national wastewater monitoring revealed that temporal shifts in extended-spectrum β-lactamase (ESBL)-producing *Escherichia coli* in wastewater suggested shifts in community carriage rates (16). Additionally, in Basel, multi-year monitoring revealed an increasing prevalence of ESBLs in hospital-associated wastewater, indicating elevated resistance within healthcare settings (17). Switzerland also maintains a long-standing clinical AMR surveillance network, with multi-year datasets documenting resistance trends in healthcare settings (18–20). While most prior work has examined wastewater and clinical datasets separately, integrating these complementary sources may offer a more holistic view of AMR dynamics in communities and clinics (21).

Given the linkages between AMR in wastewater and AMR in clinical settings, trends observed through wastewater-based surveillance may be informative of trends in clinical settings. As the proportion of resistant bacteria within a species increases in communities, as reflected in wastewater, we may expect a parallel increase in the proportion of AMR infections in clinics (8). Similarly, we may expect that as AMR increases in wastewater, there will be an increase in the consumption of antibiotics used to treat such infections. Finally, we may expect that as antibiotic consumption increases, there is increased selective pressure driving increases in AMR in the community, and therefore also in wastewater (22).

In this study, we conducted longitudinal wastewater monitoring of three clinically relevant ARB listed among the highest priority resistant pathogens by the World Health Organization: ESBL-*E. coli*, carbapenem-resistant *E. coli* (CR-*E. coli*), and vancomycin-resistant *Enterococcus faecalis/faecium* (VRE) (23). The study was conducted in Switzerland from 2021 through 2024. Using culture-based quantification and sampling on a weekly to twice-monthly basis, we tracked multi-year trends across six locations. We integrated wastewater findings with data on the proportion of resistant clinical infections and with antibiotic consumption data to investigate potential associations between community-level AMR signals in wastewater, clinical resistance patterns, and antibiotic use.

## Materials and Methods

### Experimental design

Monitoring of extended-spectrum β-lactamase-producing *Escherichia coli* (ESBL*-E. coli)* began in November 2021, as reported in Conforti et *al.* (2024), for the period from November 2021 to November 2022 (16). In November 2022, surveillance was expanded to include carbapenem-resistant *E. coli* (CR-*E. coli*) and vancomycin-resistant *Enterococcus faecium/*faecalis (VRE). This study presents data collected up to December 2024 for all antibiotic-resistant bacteria (ARB). Wastewater sampling was conducted weekly until the end of January 2024 and then every other week thereafter. Each sample was a 24-hour flow-proportional composite raw influent. Samples were collected by the wastewater treatment plant (WWTP) personnel and transported to Eawag, the Swiss Federal Institute of Aquatic Science and Technology, in Duebendorf. Samples were always kept refrigerated and processed within 48 hours of collection.

### Wastewater treatment plants

Six WWTPs across Switzerland were included in the study: ARA Altenrhein, ARA Chur, STEP d’Aïre Genève, ARA Sensetal Laupen, IDA CDA Lugano, and ARA Werdhoelzli Zurich (**Fig. 1)**. These WWTPs collectively serve approximately 1.23 million residents, corresponding to 14% of the national population (16).

**Figure 1:**
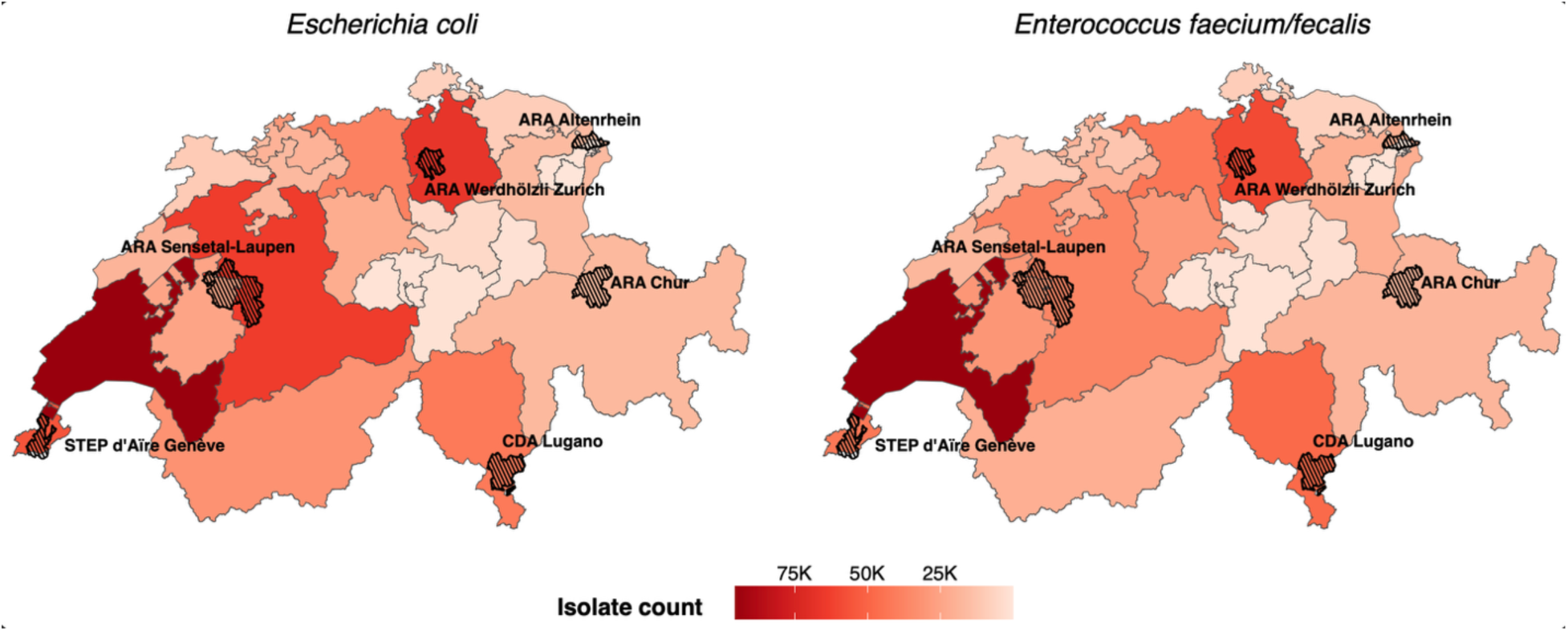
A map of Switzerland with the catchments of the wastewater treatment plants overlaid on the total number of clinical isolates screened for antimicrobial resistance by canton during the study period. The maps display the cumulative number of clinical isolates for *Escherichia coli* (left) and *Enterococcus faecium/faecalis* (right), as reported by ANRESIS, the Swiss Centre for Antibiotic Resistance (https://www.anresis.ch/). Data for *E. coli* were collected between November 2021 and December 2024, whereas data for *Enterococcus* spp. cover the same period as the corresponding wastewater sampling. The canton-level data were extracted from the national ANRESIS database, which compiles anonymized antimicrobial resistance results from over 35 human medical laboratories across Switzerland (https://www.anresis.ch/, accessed 21 March 2025). Locations of the six wastewater treatment plants included in this study are indicated with hatched areas and labeled accordingly.

### Bacterial culturing and enumeration

Chromogenic media were used for the selective enumeration of target bacteria: CHROMagar™ ESBL for ESBL-*E. coli*, CHROMagar™ mSuperCARBA™ for CR-*E. coli*, CHROMagar™ VRE for VRE, and CHROMagar™ VRE without supplement for total *Enterococcus faecium/faecalis* (CHROMagar, France). Media were prepared according to the manufacturer’s instructions. Solidified plates were stored at 4°C for up to 6 weeks until use.

For ESBL-*E. coli*, 100 µL of undiluted wastewater was plated onto CHROMagar™ ESBL. For CR-*E. coli*, 100 µL of undiluted wastewater was plated onto CHROMagar™ mSuperCARBA™. Total *E. coli* was quantified by plating serial 100-fold dilutions on CHROMagar™ Orientation. For VRE, 100 µL of undiluted sample was plated on CHROMagar™ VRE; total *Enterococcus faecium/faecalis* was enumerated using CHROMagar™ VRE without supplement with 1000-fold dilutions applied until January 2024 and 100-fold dilutions thereafter. No distinction was made between *E. faecium* and *E. faecalis*.

All samples were plated in single replicates until 8 February 2022 and in duplicates thereafter. Plates were incubated at 37°C for 24 hours. Colonies were identified based on their characteristic morphology as specified by the manufacturer: dark pink to reddish for *E. coli*, ESBL-and CR-*E. coli,* and pink for VRE and total *E. faecium/faecalis*.

### Comparison with resistance data from clinical settings

To provide context for the wastewater data, data from clinical settings covering the same period were obtained from ANRESIS, the Swiss Centre for Antibiotic Resistance, which provided antibiotic resistance data from 35 human medical laboratories (https://www.anresis.ch/, accessed on the 21 March 2025). The data provided were daily and stratified by Swiss cantons, including information on the selected ARB target, along with their susceptibility and resistance profiles.

### Comparison with antibiotic consumption data

To contextualize resistance patterns observed in wastewater, national antibiotic consumption data were obtained from IQVIA^TM^ Switzerland, a private drug market investigation company providing an exhaustive dataset of antibiotic consumption (corresponding to sales data from pharmaceutical industries to public pharmacies, self-dispensing physicians, and hospitals). Monthly antibiotic use between November 2021 and December 2024 was provided, reported at the substance level. Consumption was expressed as Defined Daily Doses per 1,000 inhabitants per day (DID), in accordance with WHO guidelines, and was reported at the national level. The dataset was restricted to substances relevant to the resistance phenotypes monitored in this study (**Table S1**). Antibiotics were classified according to their role as either selection agents (those with the potential to select for resistance in the target organism) or treatment agents (those used for treating infections caused by the target organism).

### Calculations and statistical analyses

All analyses were conducted using R (v4.1.1) and R Studio (v2024.12.0+467). The proportion of ARB in each wastewater sample was expressed as a percentage and calculated by dividing the number of colony-forming units (CFUs) on selective agar with antibiotics by the number of CFUs on selective agar without antibiotics. When duplicate measurements were available, the arithmetic mean of the two replicates was used to calculate the percentage; otherwise, the single replicate was used. Similarly, for clinical ARB, the same calculation method was applied, but with counts of clinical isolates instead of CFUs, such that the proportion is estimated as the number of resistant clinical isolates over the total number of resistant and susceptible isolates. To compare wastewater and clinical compartments, data were aggregated by week and at the national level. For wastewater, samples collected every two weeks were assigned to their corresponding week, and intervening weeks without data were removed from analysis. For ARB percentages in wastewater, values were averaged across all WWTPs, accounting for the population of each catchment to ensure population-weighted averages. In contrast, for ARB percentages in clinical settings, the population was not used for normalization. The Mann-Kendall test was employed to evaluate the increase in the percentage of ARB in wastewater and clinical settings over the study period. The Sen’s slope test was computed to estimate the rate of resistance increase over time. Time series decomposition was performed to assess and separate seasonal, trend, and random components, ensuring that observed trends were not driven by seasonality. Weekly national clinical and wastewater resistance percentages were analyzed for each bacterial target to assess temporal associations between the two surveillance streams. To account for temporal autocorrelation, trend, and seasonality, the time series were first modeled using autoregressive integrated moving average (ARIMA) and exponential smoothing state-space (ETS) models, and residuals were evaluated for white noise using the Ljung–Box test. Cross-correlation analyses were then performed on the model residuals for assessing lead–lag relationships independent of internal temporal structure, utilizing the *ccf()* function in R. Bartlett’s formula was employed to derive approximate confidence intervals for cross-correlation, which were used to identify statistically significant lags (α=0.05; |r| ≥ 1.96/√n).

For comparisons with antibiotic consumption, total antibiotic use was calculated by summing inpatient and outpatient defined daily doses per 1,000 inhabitants per day (DID). As antibiotic use data were available only at the monthly level, all related analyses were performed on monthly aggregated data. For each month, the total DID corresponding to the antibiotics associated with each resistance phenotype (**Table S1**) was computed. When multiple antibiotics were relevant to a single phenotype, their DIDs were summed to obtain a cumulative value, which assumes equal effects of antibiotics on outcomes. To align with these monthly data, weekly wastewater ARB percentages were averaged within each month to match the exact temporal resolution. This monthly alignment was used for both correlation and cross-correlation analyses between antibiotic use and resistance in wastewater.

## Results and discussion

### Presence and detection of antibiotic-resistant bacteria in Swiss wastewater

A total of 772 wastewater samples were collected during the study period from November 2021 to December 2024. Of these, 764 were plated for *E. coli*, 762 for extended-spectrum β-lactamase-producing (ESBL) *E. coli*, 473 for carbapenem-resistant *E. coli* (CR-*E. coli*), 468 for *Enterococcus faecium/faecalis*, and 469 for vancomycin-resistant enterococci (VRE) (**Table S2, Table S3**). Most samples were processed in technical duplicates, with the percentage of samples having both replicates ranging from 91.1% for ESBL-producing *E. coli*, 91.5% for *E. coli*, 97.9% for *Enterococcus faecium/faecalis*, 99.2% for CR-*E. coli*, and 99.4% for VRE.

We observed that detection rates were high across all bacterial targets. Total *E. coli* and ESBL-*E. coli* were detected in 100% of tested samples, *Enterococcus faecium/faecalis* and VRE were detected in 99.6% and 97.9% of tested samples, respectively, while CR-*E. coli* was detected in 80.8% (**Table S2**). These frequencies indicate that all ARB targets were widespread and stably present in the sampled communities, with the lower detection rate for CR-*E. coli* likely reflecting its lower prevalence, reduced shedding amongst carriers, or reduced environmental persistence during sewer transport and sample storage (24).

### ARB percentage in wastewater

To assess national trends in the proportion of ARB in wastewater, resistance percentages were calculated for each sample and aggregated weekly using population-weighted means. These national-level percentages were then summarized across the study period. The median percentage of ESBL-*E. coli* was 2.0% (interquartile range [IQR] 0.8%), followed by 0.2% (IQR 0.3%) for VRE, and 0.1% (IQR 0.1%) for CR-*E. coli* (**Table 1**). The proportions of CR-*E. coli* and VRE were lower than those of ESBL-*E. coli*, suggesting either reduced carriage within the population, differences in shedding dynamics, or reduced persistence in sewers or during sample transport. While ESBL-*E. coli* represented the most prevalent target in our study, its proportion remained lower than that reported in urban wastewater from Norway and Sweden (25,26), which used culture-and isolate-based screening methods different from ours.

**Table 1:**
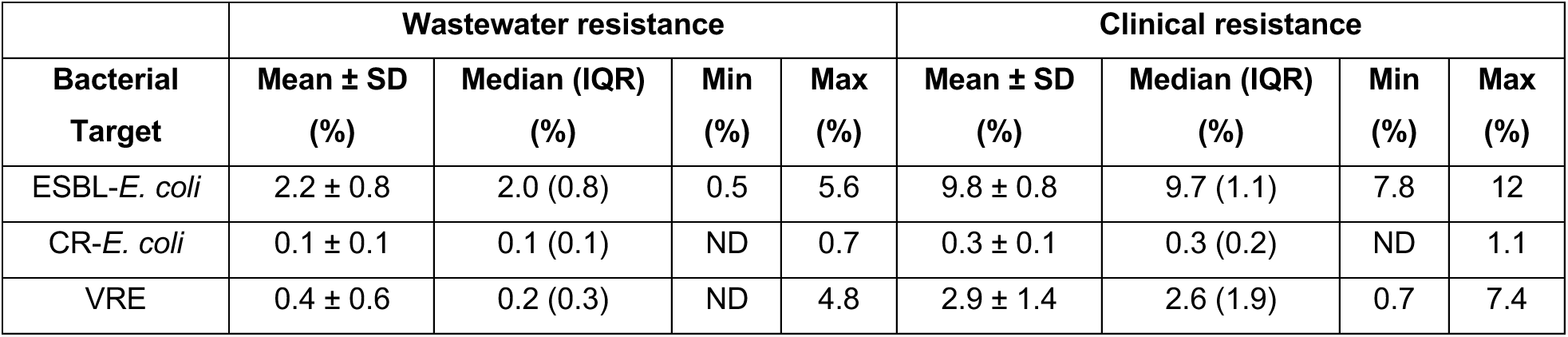
Comparison of weekly resistance percentages for each antimicrobial-resistant bacterium (ARB) between wastewater and clinical data. Values represent the mean ± standard deviation (SD), median with interquartile range (IQR), and range (minimum–maximum) across the national study period. ARBs include ESBL-producing *E. coli* (ESBL-*E. coli*), carbapenem-resistant *E. coli* (CR-E. coli), and vancomycin-resistant *Enterococcus faecium/faecalis* (VRE). In wastewater data, ND indicates that the bacteria were not detected in a sample. Non-detects were replaced with zero for the calculation of the mean and standard deviation.

Over the study period, we observed a moderate monotonic increase in the percentage for ESBL-*E. coli* (Mann-Kendall test, 𝜏 = 0.38, *p* < 0.001) and VRE (𝜏 = 0.35, *p* < 0.001) in wastewater at the national level. The observed increase corresponds to a 0.5% (±0.1%) annual increase for ESBL-*E. coli*, and of a 0.2% (±0.1%) annual increase for VRE (**Table S4**). Time series decomposition confirmed that these upward trends were not driven by seasonal variation, as the seasonal component remained stable over time, while the trend component showed a consistent increase (**Fig. S1**). Based on the conceptual model established in Conforti et al. (2024), which suggests that the proportion of resistance in a community is influenced by the prevalence and the average proportion of resistance within a single carrier (16), these increasing trends suggest a growing burden of ESBL-*E. coli* and VRE carriage in the population over the study period. However, the trends could also be influenced by factors such as gradual shifts in wastewater inputs (e.g., from industrial or agricultural sources). In contrast, no significant trend was detected for CR-*E. coli* (𝜏 = 0.02, *p* = 0.75) (**Fig. 2, TableS4**). We interpret the absence of trends in CR-*E. coli* as indicating stable carriage over time, such as would be observed if there was a consistent proportion of the population shedding into the wastewater who are carriers of CR-*E. coli*.

**Figure 2:**
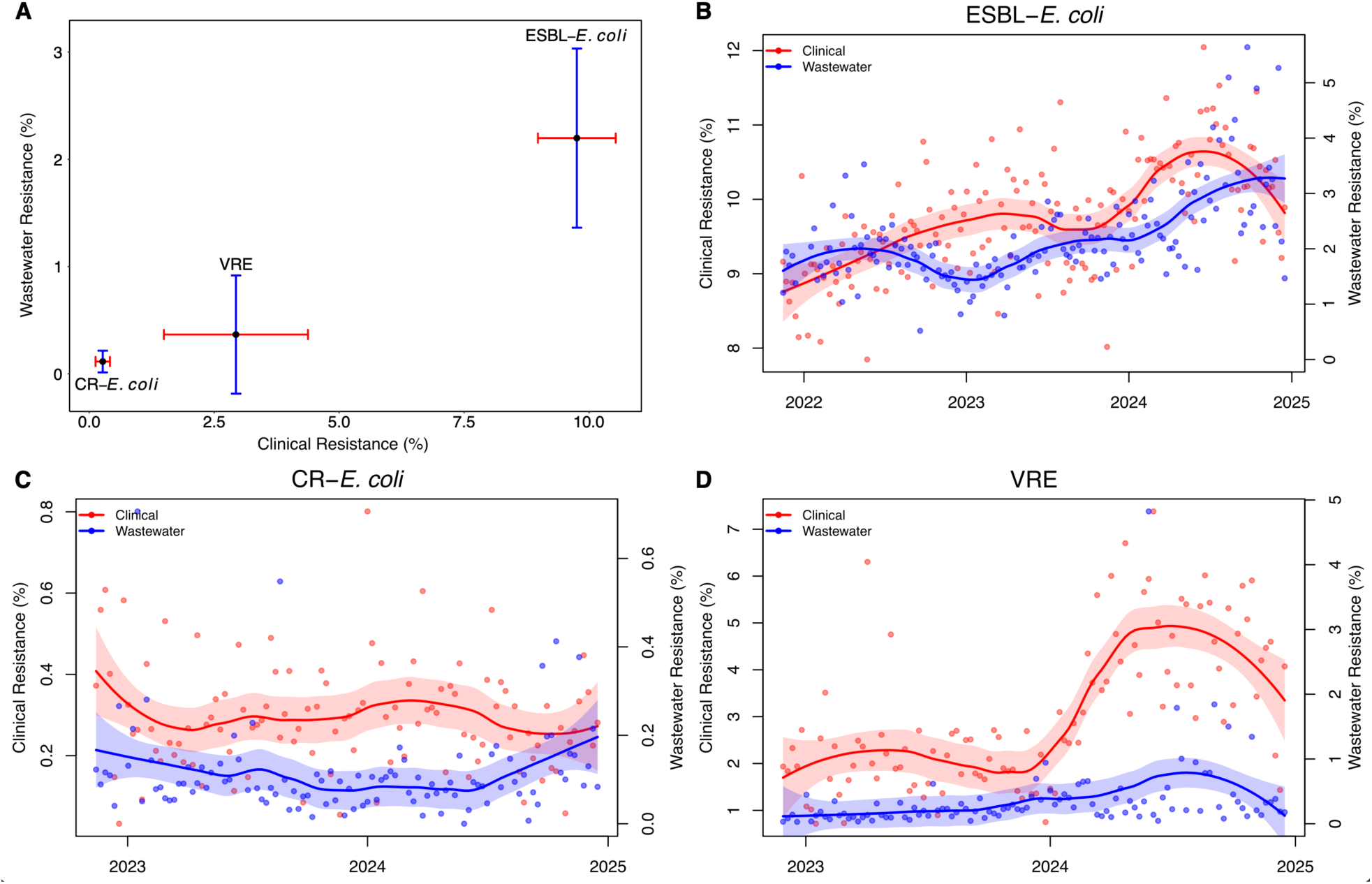
Temporal trends in clinical and wastewater resistance for three antibiotic-resistant bacterial targets (2021–2024). (A) Comparison of mean proportions of antibiotic-resistant bacteria in wastewater and clinical settings in Switzerland. Each point represents the average resistance percentage for ESBL–*E. coli,* CR–*E. coli,* and VRE, with error bars showing ±1 standard deviation. Data for ESBL–*E. coli* cover 2021–2024, whereas data for CR–*E. coli* and VRE cover 2022–2024. (B-D) Weekly clinical (red) and wastewater (blue) resistance percentages over time for ESBL-*E. coli*, CR-*E. coli*, and VRE. Clinical data represent the proportion of resistant isolates among all isolates reported to ANRESIS. Wastewater data represent the percentage of resistant colonies among all colonies plated from each sample, aggregated nationally and weighted by catchment population.

When focusing on the six WWTPs rather than the national level, we observed significant differences in resistance percentages between WWTPs for ESBL-*E. coli*, CR-*E. coli*, and VRE (Kruskal-Wallis *p* < 0.001) (**Fig. S2**, **Table S5**). These differences could be influenced by local antibiotic use, healthcare settings, demographic factors, or differences in fate and transport of AMR within the sewer network (27). Importantly, across all three targets, pairwise comparisons did not identify any WWTP as persistently higher or lower than others (Dunn’s test, **Table S5**), indicating that observed national resistance trends are not driven by just one or a few WWTPs.

Temporal trend analysis at the WWTP level revealed significant increases in ESBL-*E. coli* percentages at five out of the six WWTPs (Altenrhein, Chur, Geneva, Laupen, and Zurich), with no trend observed in Lugano. VRE increased significantly at all WWTPs, while CR-*E. coli* trends varied, with significant decreases in Altenrhein and Chur (**Table S6**).

### ARB percentage in clinical settings

To complement the interpretation of ARB percentages in wastewater, data on clinical isolates were obtained from ANRESIS, covering the period from 2021 to 2024. The dataset comprised human isolates collected across Switzerland and stratified by canton. Over the entire study period, a total of 601,306 *E. coli* and 90,468 *Enterococcus faecium/faecalis* isolates were reported (**Fig. 1, Table S7**). As expected, variability in isolate counts was observed across cantons (**Fig. 1, Fig. S3**). Indeed, a strong positive association was found between cantonal population size and the number of clinical isolates, with Spearman correlation coefficients of 0.90 for *E. coli* and 0.93 for *Enterococcus faecium/faecalis* (**Fig. S3**). Of the total clinical isolates, 9.7% (n = 58,498) were identified as ESBL-*E. coli*, 0.2% (n = 1,450) as CR-*E. coli*, and 2.9% (n = 2,639) as vancomycin-resistant *Enterococcus faecium/faecalis* (VRE) (**Table S7**).

National resistance percentages were calculated weekly and summarized across the study period. The median percentage of ESBL-*E. coli* was 9.7% (IQR 1.1%), followed by 2.6% (IQR 1.9%) for VRE, and 0.3% (IQR 0.2%) for CR-*E. coli* (**Table 1**).

Amongst the clinical surveillance data, a moderate monotonic increase was detected for ESBL-*E. coli* over time (Mann-Kendall test, 𝜏 = 0.44, p < 0.001), while weaker but significant increases were observed for CR-*E. coli* (𝜏 = 0.19, p < 0.001) and VRE (𝜏 = 0.22, p < 0.001) (**Table S4, Fig. 2**). The observed increase corresponds to a 0.5% (±0.1%) annual increase for ESBL-*E. coli*, 0.5% (±0.2%) annual increase for VRE, and 0.04% (±0.02%) annual increase for CR-*E. coli* (**Table S4**). Time series decomposition confirmed that the upward trends were independent of seasonality, with a stable seasonal component and a consistently increasing trend (**Fig. S4**). These observed trends reflect growing resistance in healthcare settings and, notably, align with wastewater data for ESBL-*E. coli* and VRE.

### Comparison of ARB percentages in wastewater and in clinical settings

The rank order of resistance proportions in clinical data (ESBL-*E. coli* > VRE > CR-*E. coli*) mirrors that observed in wastewater (**Fig. 2**), suggesting that the percentages of AMR in the community are linked to those observed in clinical settings. Notably, ARB percentages were consistently higher in clinical settings than the percentages in wastewater for all ARB (**Table 1, Fig. S2**). The observed difference in values reflects that the two data sources represent different proportions: the proportion of resistance in clinical settings reflects the share of all resistant infections, whereas for wastewater, the proportion reflects the share of resistant bacteria amongst all (resistant and susceptible) bacteria. This discrepancy reflects the fundamentally different nature of the two data sources, where wastewater captures pooled contributions from the entire population, including healthy individuals who may carry ARB asymptomatically or at lower levels. In contrast, clinical data are often biased toward individuals seeking medical attention, which includes hospitalization as well as targeted screening. These factors can lead to higher resistance prevalence in clinical settings as compared to community settings (16,29). If we assume that wastewater indicates the overall proportion of AMR bacteria circulating in the population, then higher proportions of ARB observed in clinical settings compared to wastewater may suggest two things. First, susceptible bacteria are less likely to cause an infection and therefore are less likely to be sampled. If infections caused by susceptible bacteria occur, they may be mild, self-resolving, and less likely to be diagnosed or sampled in clinics. Second, resistant bacteria may be more likely to cause an infection requiring medical attention, making them more likely to be diagnosed, reported, and sampled.

The relationship in trends of ARB percentages in wastewater and clinical settings was further investigated on a weekly basis. Spearman correlation analysis between percentages in wastewater and clinical settings indicated a moderate positive correlation for ESBL-*E. coli* (r = 0.36, p < 0.001), a weak, but not significant, positive correlation for VRE (r = 0.19, p = 0.06), and no correlation for CR-*E. coli* (r = –0.05, p = 0.625) (**Fig. S5**). The moderate correlation for ESBL-*E. coli* suggests that temporal trends are shared between clinical and environmental compartments. The observed increase over time was consistent for wastewater and clinical resistance data, both showing a 0.5% annual increase in ESBL-*E. coli*. In contrast, the increase in VRE was higher in clinical resistance data (+0.5%) compared to wastewater resistance (+0.2%) annually. While previous cross-sectional studies reported correlations between clinical resistance and resistance detected in wastewater *E. coli* (11,30), these were based on single time points per country and did not capture the temporal dynamics. Our longitudinal analysis enabled a comparison of temporal relationships between clinics and wastewater within a single location (16,29). This trend was not observed, however, for CR*–E. coli*, which highlights a decoupling between the proportion of resistant *E. coli* in clinical infections and the proportion detected in wastewater. In contrast to VRE and ESBL*–E. coli*, the overall proportion of CR*–E. coli* was low and showed the smallest range of values across the study period. We note that the low proportion is linked to low CR-*E. coli* concentrations in wastewater, with counts of CR*–E. coli* often below 10 CFU per plate (and occasionally close to the lower limit of reliable quantification), where stochastic variability is high. Variation in stochasticity may have obscured temporal trends. Analyzing volumes larger than 100 µl, such as through sample concentration, may reduce variability in estimates of the proportion of CR-*E. coli* amongst total *E. coli*.

Temporal links between clinical and wastewater resistance trends were weak and inconsistent. Cross-correlation analysis using de-autocorrelated residuals showed short and weak lead-lag relationships between clinical and wastewater resistance trends (**Table 2**, **Fig. 2, Fig. S6**). For ESBL-*E. coli*, the peak correlation (r = −0.17) was observed when clinical trends slightly preceded wastewater trends by one week. For CR-*E. coli*, the peak correlation (r = −0.18) was observed at a lag of −5 weeks, though no Bartlett-significant range was identified, indicating a weak and unstable association. For VRE, the peak correlation (r = −0.26) occurred when wastewater trends preceded clinical trends by four weeks, suggesting a modest lead of wastewater over clinical resistance. All correlations were negative, and their small magnitudes indicate that increases in clinical resistance were not associated with corresponding increases in wastewater resistance. Overall, these results suggest that there is no consistent or meaningful temporal relationship between clinical and wastewater resistance once autocorrelation and seasonality are accounted for, implying that the temporal dynamics in the two surveillance streams were largely independent over the two years of data analyzed here.

**Table 2:** Cross-correlation between residuals of clinical and wastewater ARB percentages by bacterial target. The Peak Lag column shows the single lag (in weeks) with the highest absolute correlation between the residuals of clinical and wastewater time series (positive lags indicate wastewater leads; negative lags indicate clinical leads). The Significant Range indicates the contiguous set of lags that are statistically significant according to Bartlett’s approximation (α = 0.05; |r| ≥ 1.96/√n) and have the same sign as the peak correlation. The Max Correlation (r) represents the magnitude of the cross-correlation at the peak lag, and the Lag Interpretation describes the direction of the relationship.

| Bacterial Target | Peak Lag (weeks) | Significant Range (weeks)<sup>a</sup> | Max Correlation (r) | Lag Interpretation |
| --- | --- | --- | --- | --- |
| ESBL-*E. coli* | -1 | -1 • -1 | -0.17 | Clinical leads wastewater |
| CR-*E. coli* | -5 | - | -0.18 | Clinical leads wastewater |
| VRE | 4 | 4 • 4 | -0.26 | Wastewater leads clinical |

<sup>a</sup> A dash (–) in the ‘Significant Range’ column indicates that the peak lag did not meet the criteria for a contiguous significant range. Significance was assessed using the Bartlett 95% confidence band, which requires the correlation at a given lag to exceed $\pm 1.96/\sqrt{n}$ (where $n$ is the number of paired time points) and to have the same sign as the peak correlation. If no contiguous block of lags satisfied both conditions, the significant range is reported as “–”.

### Comparison of ARB percentages in wastewater and antibiotic consumption

ARB percentages in wastewater were compared to antibiotic consumption under two regimes: first, wastewater percentages were compared to consumption of antibiotics used to treat infections, and second, to consumption of antibiotics that may select for resistance. Correlations were assessed using data aggregated to the monthly level across resistance phenotypes (**Fig. 3**). At the monthly level, Spearman correlation analyses revealed weak or no associations between antibiotic consumption for treatment and ARB percentages in wastewater, with variability across resistance phenotypes (**Fig. S7**). For antibiotics selecting for resistance, ESBL-*E. coli* showed weak negative correlations (Spearman ρ = –0.36, p = 0.03), indicating that higher antibiotic consumption was associated with lower percentages of ESBL-*E. coli* in wastewater at the same time point. This suggests that increases in antibiotic consumption did not correspond to concurrent increases in wastewater resistance. Such a pattern may reflect that the study duration is insufficient to observe meaningful effects, there are delayed effects of selective pressure, or there are confounding factors, such as variable shedding rates or environmental persistence. No significant correlations were observed between CR-*E. coli* and VRE and the antibiotics selecting for their resistance (CR-*E. coli*: Spearman’s ρ = –0.21, *p* = 0.3; VRE: Spearman’s ρ = –0.05, *p* = 0.8).

**Figure 3:**
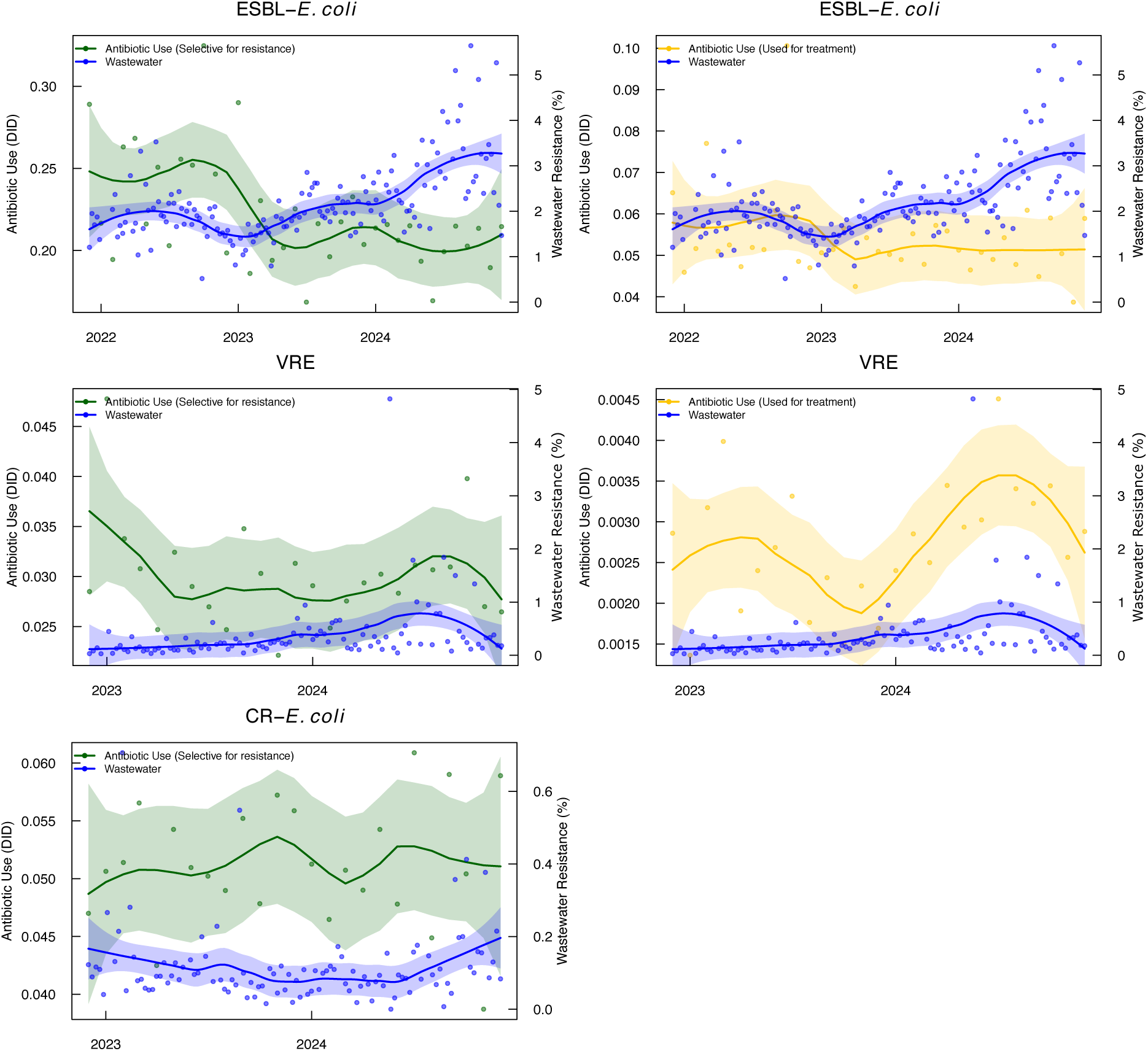
T**e**mporal **trends in wastewater resistance and antibiotic use for three antibiotic-resistant bacterial targets (2021–2024).** Left column: monthly trends in antibiotic consumption that select for resistance in the corresponding targets (green) and wastewater resistance (blue). Right column: monthly trends in antibiotic consumption that are used for treating the corresponding target (orange) and wastewater resistance (blue). Antibiotic use is expressed as defined daily doses per 1,000 inhabitants per day (DID), calculated by summing inpatient and outpatient consumption of all antibiotics relevant to each resistance phenotype (**Table S1**). Shaded bands represent smoothed trends with 95% confidence intervals. We note no analyses of CR-*E. coli* with antibiotics used for treatment because such drugs are not included in the available dataset.

For antibiotics primarily used in treatment, no meaningful correlations were observed for ESBL-*E. coli* and VRE at the monthly scale (Spearman |ρ| ≤ 0.33, p > 0.10 for both ARB). This indicates that temporal fluctuations in the use of these treatment antibiotics were not associated with concurrent changes in the proportion of corresponding ARB in wastewater. This finding is expected, as treatment antibiotics are typically prescribed in response to ongoing infections rather than driving the selection of resistance at the community level. For CR-*E. coli*, no corresponding analysis was performed because the antibiotic consumption dataset does not include drugs used specifically for the treatment of CR-*E. coli*.

Cross-correlation analyses of de-autocorrelated residuals revealed weak to moderate and inconsistent temporal associations between antibiotic use and wastewater resistance (**Table 3, Fig. S8**). For selection antibiotics, the strongest association was observed for ESBL-*E. coli*, where increases in wastewater resistance were followed two months later by a decrease in the use of antibiotics that can select for ESBL resistance (e.g., third-and fourth-generation cephalosporins; r = –0.39). This relationship was supported by a narrow Bartlett-significant range centered at a two-month lag. For CR-*E. coli*, the higher use of antibiotics that select for carbapenem resistance preceded a modest increase in wastewater resistance by approximately 11 months (r = 0.36). For VRE, greater use of vancomycin was followed by a slight rise in wastewater resistance after five months (r = 0.33).

**Table 3:** Cross-correlation between residuals of national antibiotic consumption and wastewater percentages of antibiotic-resistant bacteria (ARB) in Switzerland (2021–2024), stratified by antibiotic category (“Selection” vs “Treatment”). Selection antibiotics are those with the potential to select for the corresponding resistance phenotype. In contrast, treatment antibiotics are typically used in targeted therapy for infections caused by the corresponding resistant bacteria. Correlations were computed using residuals obtained after accounting for temporal autocorrelation through ARIMA modeling. The Peak Lag column shows the single lag (in months) with the highest absolute correlation (positive lags indicate wastewater leads; negative lags indicate antibiotic use leads). The Significant Range lists contiguous Bartlett-significant lags (α≈0.05; |r| ≥ 1.96/√n).

| Bacterial Target | Antibiotic Category | Peak Lag (months) | Significant Range (months)<sup>a</sup> | Max Correlation (r) | Lag Interpretation |
| --- | --- | --- | --- | --- | --- |
| CR-*E. coli* | Selection | -11 | - | 0.36 | Antibiotic use leads wastewater |
| ESBL-*E. coli* | Selection | 2 | 2•2 | -0.39 | Wastewater leads antibiotic use |
| VRE | Selection | -5 | - | 0.33 | Antibiotic use leads wastewater |
| ESBL-*E. coli* | Treatment | 2 | - | -0.29 | Wastewater leads antibiotic use |
| VRE | Treatment | -2 | - | 0.37 | Antibiotic use leads wastewater |

<sup>a</sup> A dash (–) in the ‘Significant Range’ column indicates that the peak lag did not meet the criteria for a contiguous significant range. Significance was assessed using the Bartlett 95% confidence band, which requires the correlation at a given lag to exceed $\pm 1.96/\sqrt{n}$ (where $n$ is the number of paired time points) and to have the same sign as the peak correlation. If no contiguous block of lags satisfied both conditions, the significant range is reported as “–”.

For treatment antibiotics, associations were similarly weak and inconsistent. For ESBL-*E. coli*, increases in wastewater resistance slightly preceded a reduction in carbapenem use by about two months (r = –0.29). For VRE, higher wastewater resistance was followed two months later by an increase in the use of linezolid (r = 0.37). Except for the ESBL-*E. coli* selection relationship, none of these correlations were Bartlett-significant, indicating that they were not consistent across neighbouring lags. Collectively, these results suggest that, once temporal autocorrelation and shared trends are removed, antibiotic consumption and wastewater resistance vary largely independently, with no substantial or systematic evidence of short-term coupling between the two. Overall, we note that our observations are correlative, not causative, so significant and meaningful correlations may be driven by hidden or unobserved factors that influence shifts in the proportion of AMR in clinics, wastewater, and in the consumption of antibiotics, both those that select for resistance and those used to treat corresponding resistance.

### Future implications and conclusions

Our findings demonstrate that wastewater-based surveillance (WBS) offers a valuable but complex lens for monitoring AMR, which can complement clinical reporting. Both wastewater and clinical datasets showed gradual increases over the study period. For CR-*E. coli* and VRE, the estimated annual percentage increase was higher in clinical resistance data than in wastewater, suggesting that increases in clinical infections with resistant bacteria outpaced changes in the proportion of resistant bacteria detected in wastewater during the study period. In contrast, ESBL-*E. coli* showed the same estimated annual increase of 0.5% in both wastewater and clinical data, indicating more closely aligned trends between the two surveillance streams. Furthermore, the rank order in the percentages of antibiotic-resistant bacteria studied was conserved across both sources (ESBL-*E. coli* > VRE > CR-E. *coli*), indicating that the proportion of resistant bacteria in the community is reflected in the proportion of resistance in clinics, and that long-term trends in wastewater could be informative beyond our study period. The lead-lag relationships between WBS and clinical data were weak and varied across ARB, underscoring the target-specific nature of these associations and suggesting that short-term trends in wastewater (week-to-week or month-to-month) are less informative. Indeed, interpretation of WBS remains complicated by factors such as non-human inputs, potentially variable environmental persistence of resistance determinants, and fate and transport processes in sewer systems, which may decouple wastewater signals from community or clinical trends. After correcting for autocorrelation and seasonality, however, cross-correlations between compartments and with antibiotic use were weak and inconsistent, implying that short-term clinical or prescribing fluctuations do not directly translate to wastewater signals. Instead, WBS primarily reflects the underlying community carriage of resistant organisms, integrating inputs from both symptomatic and asymptomatic individuals.

## Data availability

All scripts used for data analysis and figure creation, along with wastewater data, are available at https://github.com/EawagPHH/AMR_wastewater_clinical.git. Anonymized clinical resistance data from ANRESIS (https://www.anresis.ch) can be obtained upon request from ANRESIS or the corresponding author. Antibiotic consumption data may also be available upon request from the corresponding author.

## Author contribution statement

The author contributions below are according to the CRediT statement. S.C.: Conceptualization, Methodology, Formal analysis, Investigation, Data Curation, Visualization, Writing-Original Draft, Writing - Review & Editing, Visualization. M.P.: Conceptualization, Methodology, Formal analysis, Investigation, Data Curation, Visualization, Writing-Original Draft, Writing - Review & Editing, Visualization. P.S.: Methodology, Formal analysis, Writing-Review & Editing. A.W.: Data Curation, Writing - Review & Editing. C.P.: Data Curation, Writing - Review & Editing. A.K.: Conceptualization, Data Curation, Writing - Review & Editing. T.R.J.: Conceptualization, Methodology, Resources, Writing – Review & Editing, Supervision, Project administration, Funding acquisition.

## Acknowledgements

We thank the Wastewater-based Infectious Disease Surveillance (WISE) group for insightful discussions and the many past and present members of the wastewater monitoring at Eawag who assisted with plating and enumeration. We are also grateful to the staff of IDA CDA Lugano (Ticino), ARA Werdhölzli (Zurich), ARA Chur (Graubünden), ARA Sensetal Laupen (Bern), and STEP d’Aïre Genève (Geneva) for providing wastewater samples. We thank all participating ANRESIS laboratories (www.anresis.ch) and IQVIA^TM^ for providing resistance and antibiotic consumption data.

## Generative AI usage Statement

Generative AI tools (ChatGPT, OpenAI) were used solely to support coding for data analysis. No AI tools were used to generate scientific interpretations, text, or conclusions. All code and outputs were reviewed and validated by the authors.

