## Supporting Information for "Integrated analyses of longitudinal trends of antibiotic-resistant bacteria in wastewater, clinical resistance data, and antibiotic consumption in Switzerland"

**Supplementary Figures**

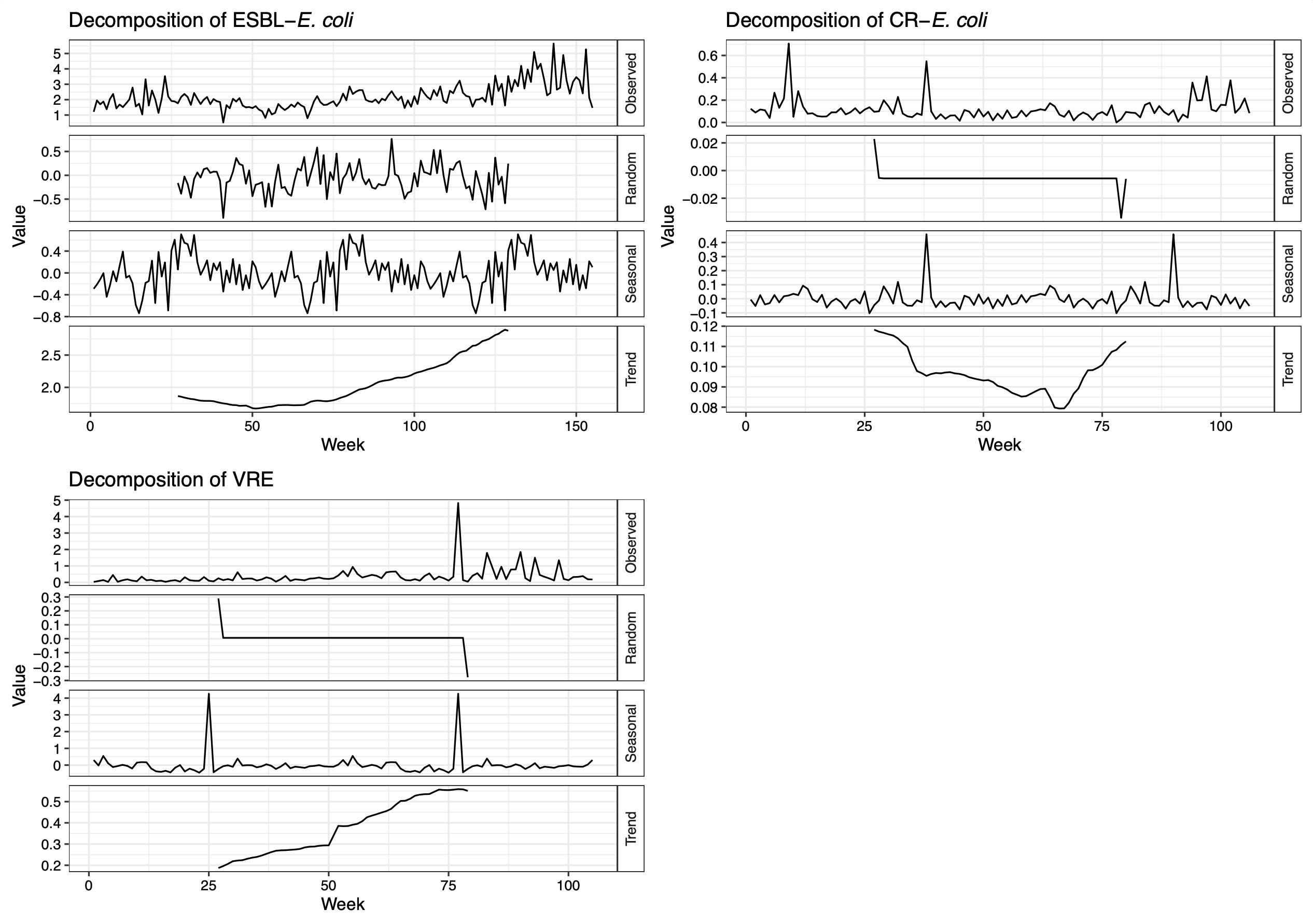

**Fig. S1**: **Decomposition analysis of wastewater resistance percentages by bacterial target.** This figure shows the decomposition of wastewater resistance percentages for three bacterial targets: ESBL-E. coli, CR-E. coli, and VRE. For each target, the weekly time series is separated into four components: the observed values represent the raw weekly resistance percentages; the trend captures the long-term direction of change after accounting for random and seasonal variation; the seasonal component reflects recurring fluctuations that indicate potential seasonal effects; and the random (residual) component represents the remaining irregular variability unexplained by the other factors. Together, these decompositions illustrate how underlying trends and seasonal patterns contribute to temporal changes in antimicrobial resistance observed in wastewater.

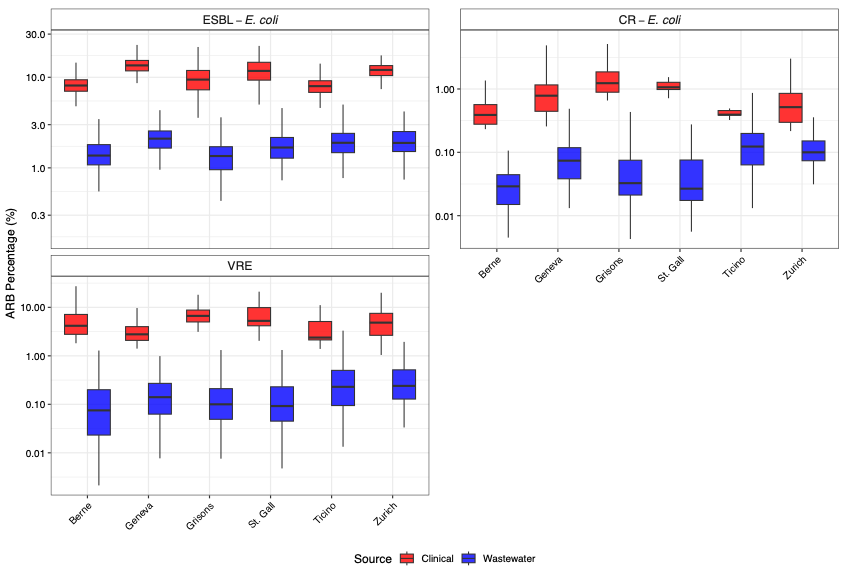

**Fig. S2**: Variation in antimicrobial-resistant bacteria (ARB) percentages across selected Swiss cantons. Boxplots show the distribution of ARB percentages in clinical and wastewater (WW) sources, plotted on a log scale. Each panel represents a different ARB type (ESBL-E. coli, CRE-E. coli, and VRE) with cantons on the x-axis and separate boxplots for each source.

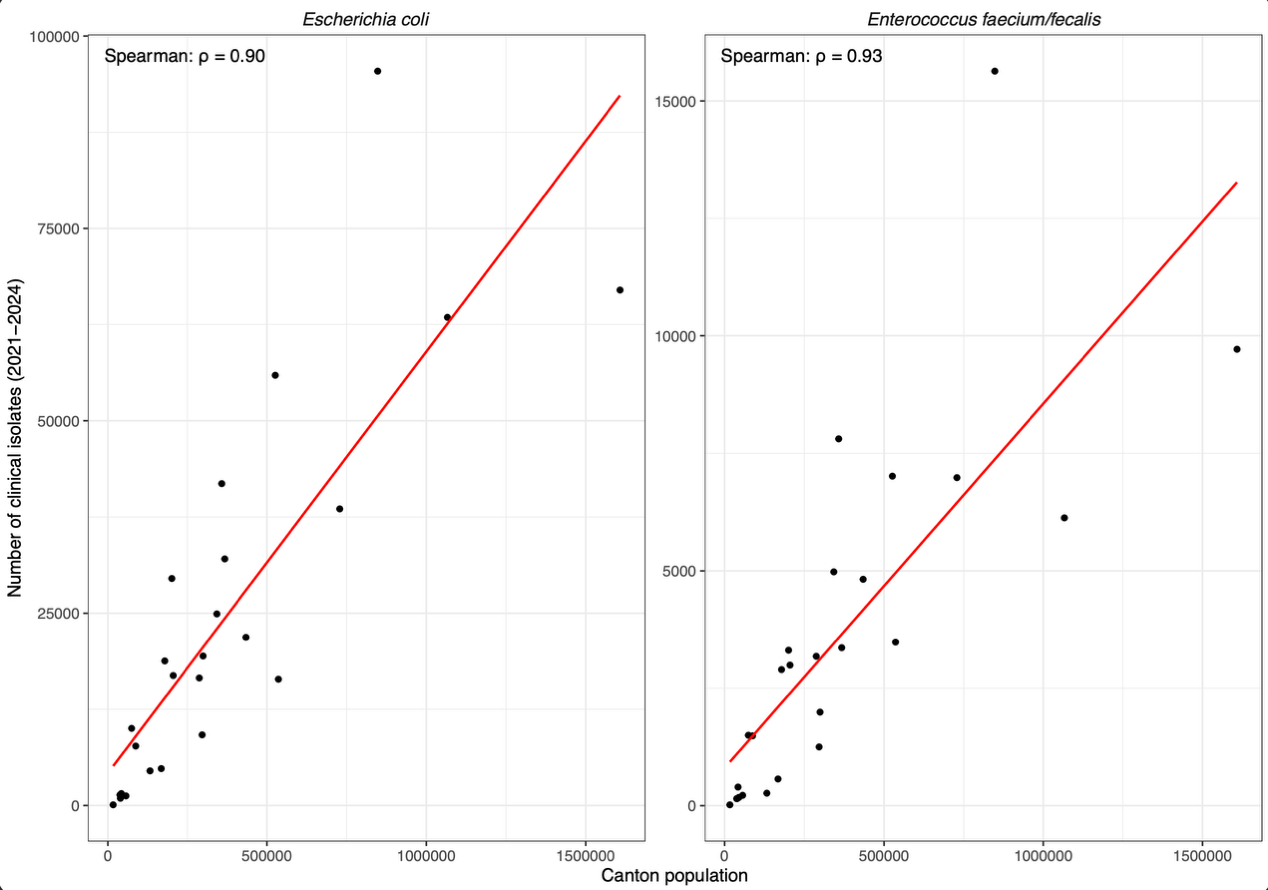

**Fig. S3**: Correlation between canton-level population size and the total number of reported clinical isolates from E. coli, and Enterococcus faecium/fecalis between 2021 and 2024. Each panel shows the linear regression (red line) and the corresponding Pearson and Spearman correlation coefficients. Data are stratified by bacterial target. Source: ANRESIS and Swiss Federal Statistical Office (population data as of 10.05.2024).

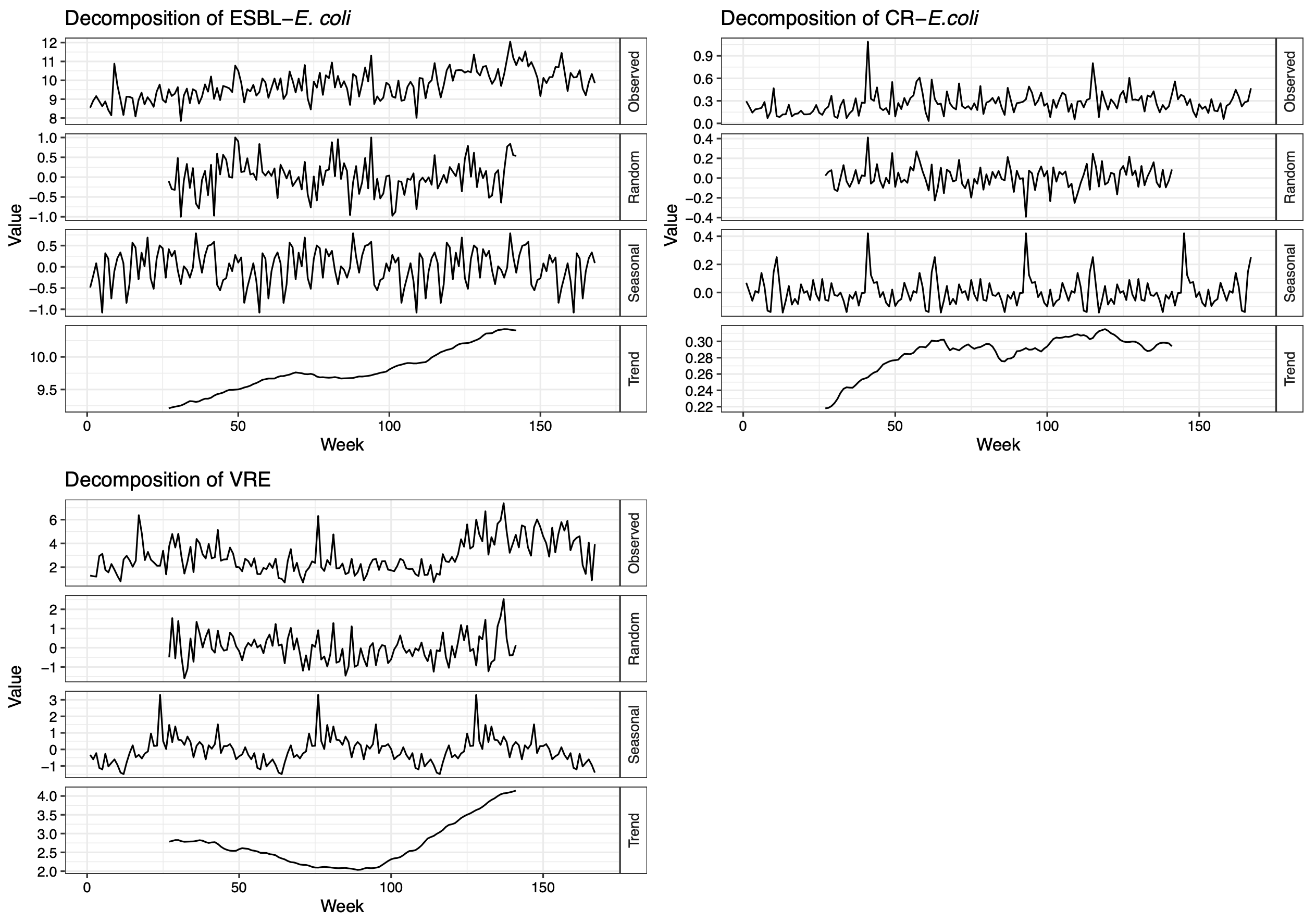

**Fig. S4**: **Decomposition analysis of clinical resistance percentages by bacterial target.** This figure shows the decomposition of clinical resistance percentages for three bacterial targets: ESBL-E. coli, CR-E. coli, and VRE. For each target, the weekly time series is separated into four components: the observed values represent the raw weekly resistance percentages in clinical isolates; the trend captures the long-term direction of change after accounting for random and seasonal variation; the seasonal component reflects recurring fluctuations that may indicate seasonal patterns in infection or reporting; and the random (residual) component represents the remaining irregular variability unexplained by the other factors. Together, these decompositions reveal how underlying trends and potential seasonal influences contribute to temporal changes in antimicrobial resistance observed in clinical settings.

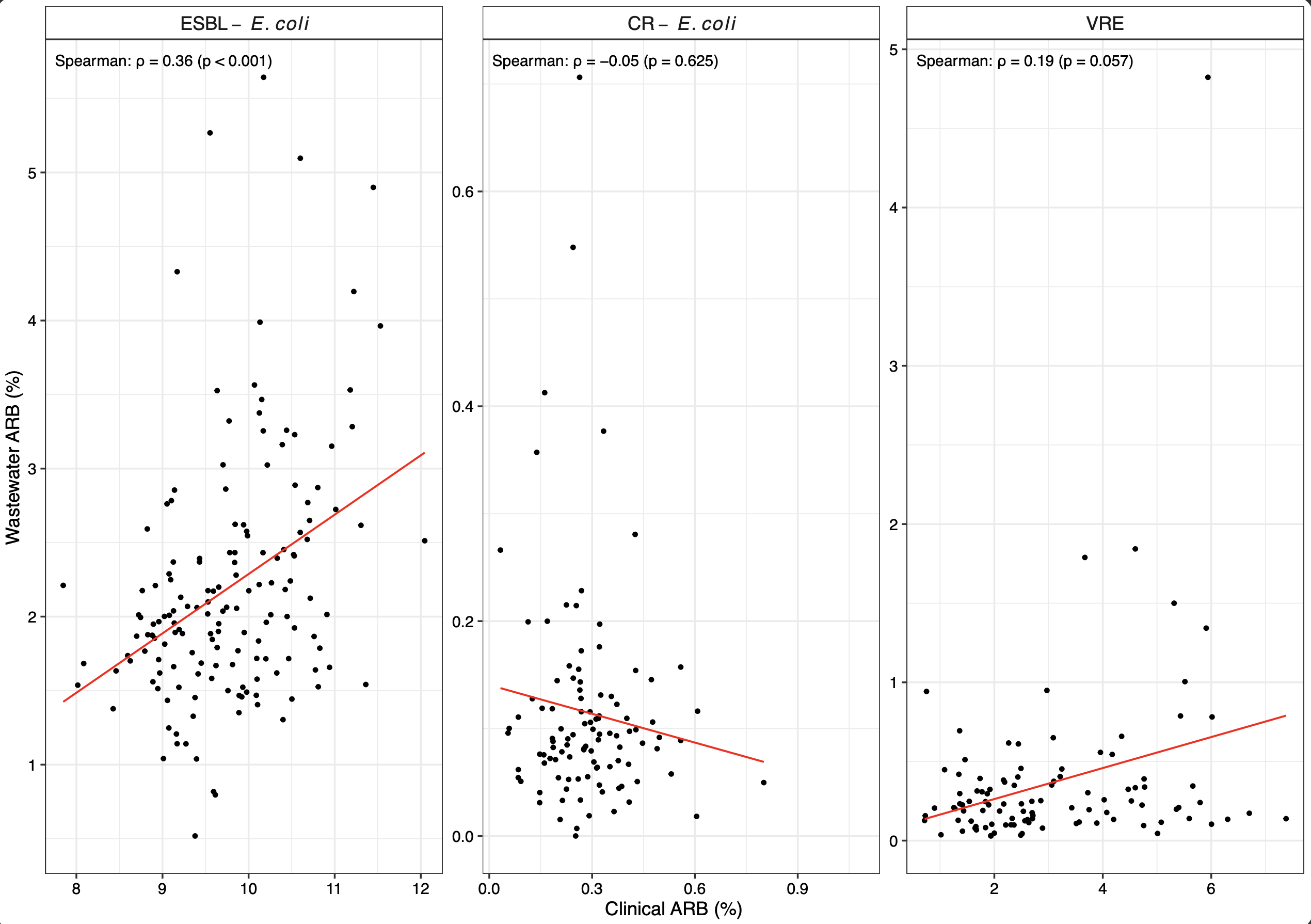

**Fig. S5**: Correlation between antibiotic-resistant bacteria (ARB) percentages in wastewater and clinical settings for three bacterial targets across weeks (2021–2024). Each point represents one week of national-level data. Spearman correlation coefficient is indicated in each panel.

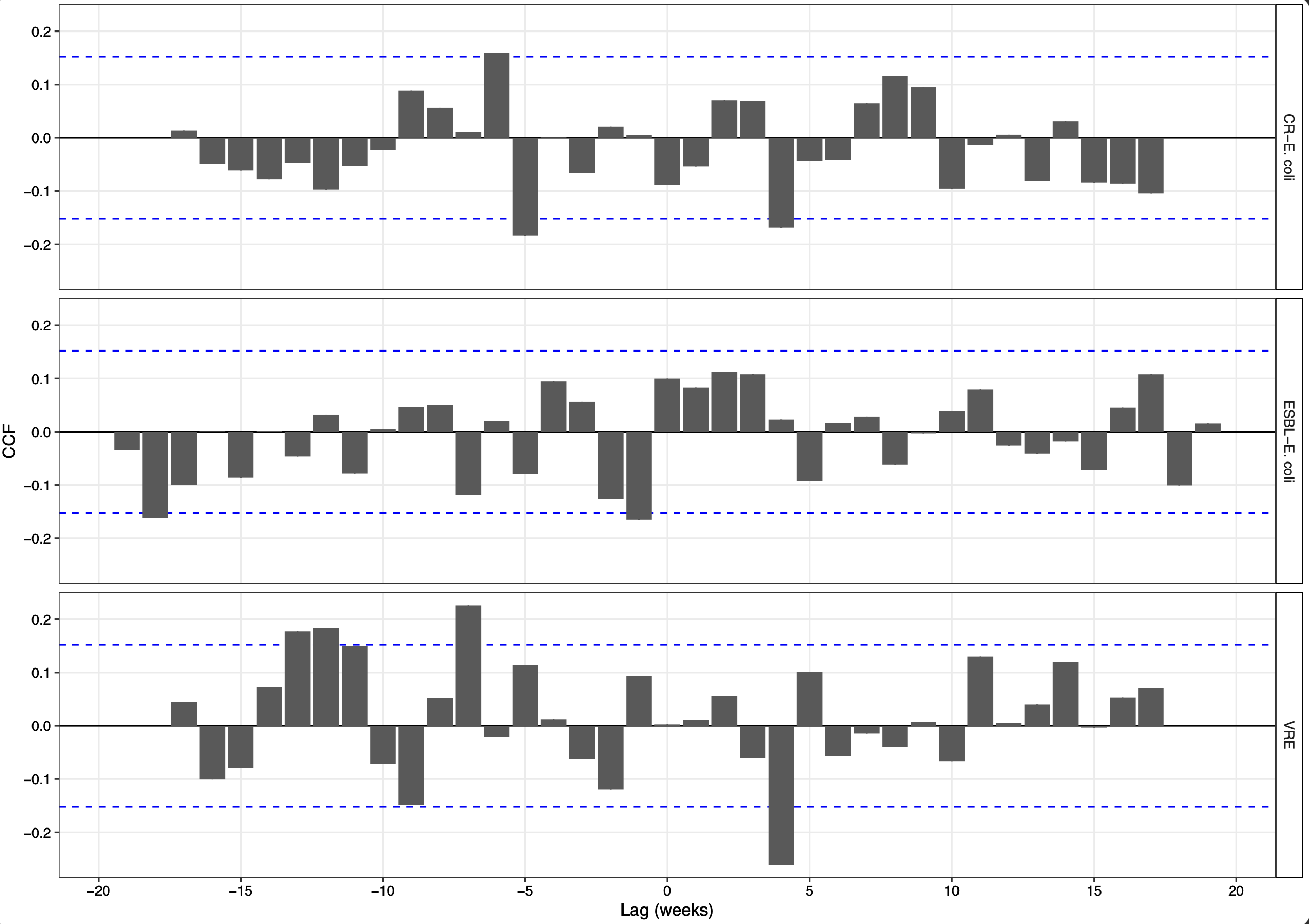

**Fig. S6**: **Cross-correlation function (CCF) between the percentage of antibiotic-resistant bacteria (ARB) in clinical and wastewater data (2021-2024).** Positive lags indicate that wastewater trends precede clinical resistance; negative lags indicate that clinical resistance precedes wastewater trends. Positive correlations indicate that the two time series move in the same direction, whereas negative correlations indicate opposite trends. Bars represent CCF values. Dashed lines show the Bartlett 95 % confidence interval (α≈0.05; |r| ≥ 1.96/√n).

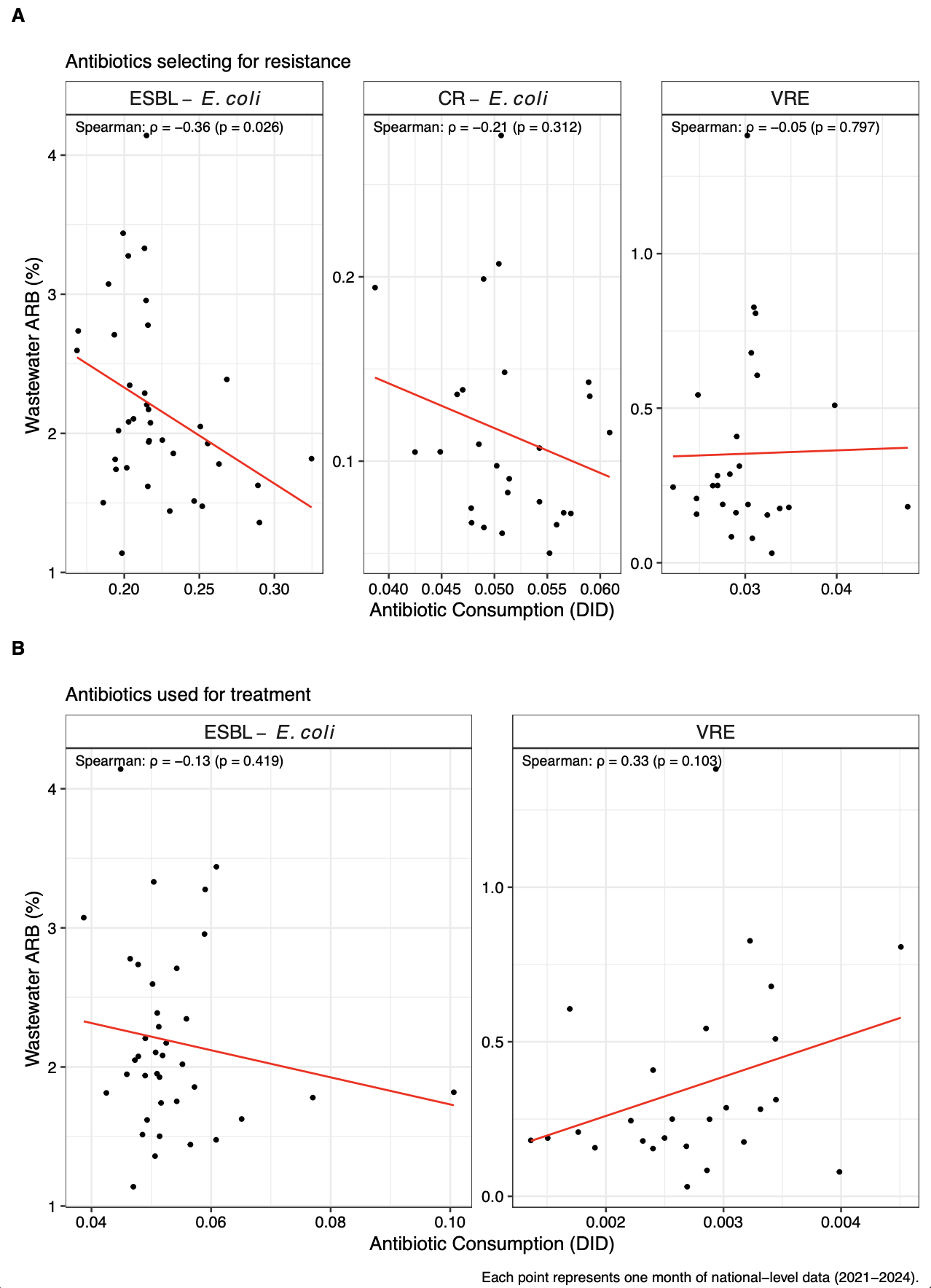

**Fig. S7**: Correlation between antibiotic consumption and wastewater percentages of antibiotic-resistant bacteria (ARB) for three bacterial targets in Switzerland, 2021–2024. Panel A shows selection antibiotics (antibiotic classes that may select for the resistance phenotype; see **Table S1** for the antibiotics included for each target), and Panel B shows treatment antibiotics. Antibiotic consumption is expressed as defined daily doses per 1,000 inhabitants per day (DID) and calculated at the national level. Wastewater ARB percentages represent the proportion of resistant colonies among all colonies plated from each sample, aggregated nationally and weighted by catchment population. Each point represents one month of data. Spearman rank correlations (ρ) with associated p-values are shown in each facet for ESBL-E. coli, CR-E. coli, and VRE.

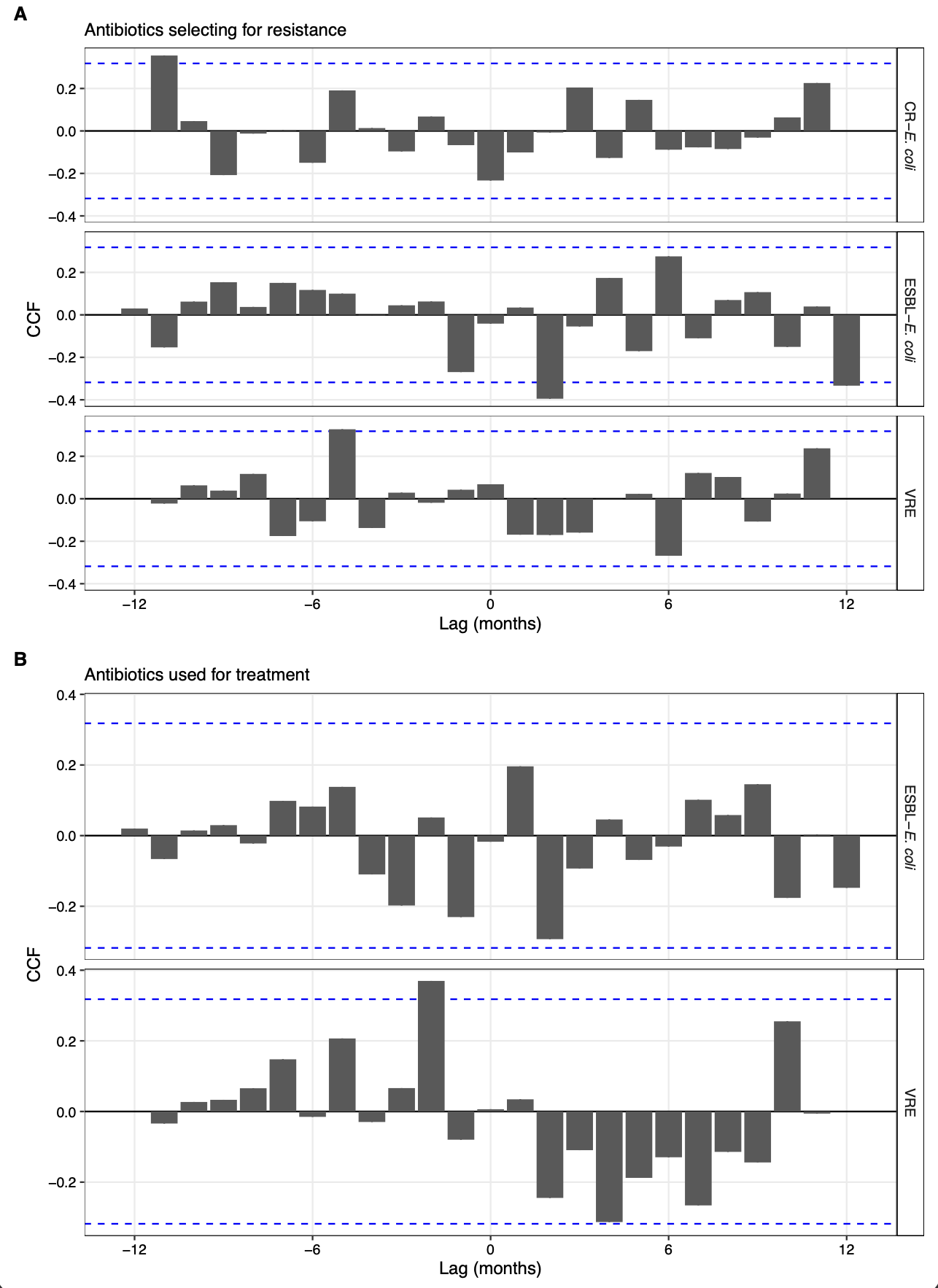

**Fig. S8**: **Cross-correlation function (CCF) between the percentage of antibiotic-resistant bacteria (ARB) in wastewater and the defined daily doses of antibiotics dispensed in the community (2021–2024).** Panel A: antibiotics selecting for resistance. Panel B: antibiotics used for treatment. Positive lags indicate that changes in antibiotic dispensing precede changes in wastewater resistance; negative lags indicate the reverse. Positive correlations indicate that the two time series move in the same direction, whereas negative correlations indicate opposite trends. Bars represent CCF values. Dashed lines show the Bartlett 95 % confidence interval (α≈0.05; |r| ≥ 1.96/√n).

**Supplementary Tables**

**Table S1**: Antibiotics included in the analysis for their potential role in the selection and/or treatment of antimicrobial-resistant bacteria (ARB) by bacterial target. The table lists each antibiotic alongside its pharmacological subclass and class, grouped by the bacterial target of interest: ESBL-*Escherichia coli*, CR-*E. coli*, and VRE. “Selection” indicates antibiotics that can drive resistance development in the target organism, while “Treatment” refers to antibiotics commonly used in clinical management of infections caused by the target.

**Table S2**: Summary of antimicrobial-resistant bacteria (ARB) detection in Swiss wastewater samples. The table reports, for each bacterial target, the number of tested samples (i.e., with at least one replicate plated), the number and percentage with both technical replicates, the number with only one replicate, and the number and percentage of samples testing positive for the target. The last row shows the total number of wastewater samples received during the study.

**Table S3**: Summary of missing ARB measurements in Swiss wastewater samples. Only dates within the defined screening period for each bacterial target were considered. Missing values are shown per location and target as comma-separated dates where both replicates were absent.

**Table S4**: Mann-Kendall trend test results for national resistance percentages in wastewater and clinical datasets. The tau coefficient (𝜏) indicates the strength and direction of monotonic trends between November 2021 and December 2024. Sen's slope values represent the estimated annual change in percentage points with 95% confidence intervals.

**Table S5**: Dunn’s multiple comparison test results for ARB percentages across WWTPs. Comparisons were adjusted using the Bonferroni method. Direction indicates whether the first WWTP in the comparison had higher or lower values.

**Table S6**: Mann-Kendall trend test results for ARB percentage trends at each WWTP. Tau values indicate the direction and strength of monotonic trends. Significance assessed at p < 0.05.

**Table S7**: Total number of clinical isolates reported to ANRESIS by bacterial target and resistance profile (2021–2024). The resistance phenotypes ESBL and CR for *E. coli*, and VRE for *Enterococcus faecium/fecalis* are shown, alongside total isolate counts and their corresponding percentages.
